# Summer heat, historic redlining, and neighborhood walking among older adults: 2017 National Household Travel Survey

**DOI:** 10.1101/2024.02.20.24303082

**Authors:** Diana Mitsova, Lilah M. Besser, Elaine T. Le

## Abstract

No known studies have examined the relationships between urban heat islands, historic redlining, and neighborhood walking in older adults. We assessed whether: 1) individual and neighborhood characteristics (including redlining score) differ by neighborhood summer land surface temperature (LST); 2) higher LST is associated with less neighborhood walking, and whether associations differ by historic redlining score; and 3) neighborhoods with discriminatory redlining scores have greater LSTs. We used data on 3,982 ≥65-year olds from the 2017 National Household Travel Survey. Multivariable negative binomial and linear regressions tested associations between LST z-score (comparing participant’s neighborhood LST to surrounding region’s LST) and self-reported neighborhood walking, and the association between living in neighborhoods redlined as “definitely declining” or “hazardous” (versus “still desirable”/“best”) and LST z-score. LSTs were higher for those in neighborhoods with higher area deprivation scores and more African American/Black residents. Older adults living in neighborhoods with higher summer LST z-scores had fewer minutes of neighborhood walking/day. This association seemed limited to individuals with neighborhood redlining scores of “still desirable”/“best”. Neighborhood redlining scores of “definitely declining” or “hazardous” (versus “still desirable” and “best”) were associated with greater neighborhood summer LSTs. Overall, these findings suggest that historically redlined neighborhoods may more often experience urban heat island effects, and older adults living in hotter neighborhoods may less often engage in neighborhood walking. Future work is needed to elucidate the impact of extreme heat on health promoting behaviors such as walking and the types of interventions that can successfully counteract negative impacts to historically disadvantaged communities.

## Introduction

Engaging in regular low-intensity physical activity, including walking, offers numerous health benefits for older adults.^1^ A vast range of observational and interventional studies consistently have documented that regular exercise helps improve cardiovascular and respiratory system function, reducing the risk of heart disease and high blood pressure.^2^ Regular moderate-intensity exercise improves the functioning of the immune system and can enhance bone density, reducing the risk of osteoporosis.^3^ Studies also suggest an association between physical activity and cell physiology linked to the genesis of cognitive decline.^1^ Exercise is also associated with mental health benefits.^4,5^ Recent meta-analyses provided evidence that walking can effectively reduce the symptoms of anxiety and depression and improve sleep health.^1^ In addition, exercise in a group setting can provide opportunities for social interaction, easing feelings of isolation and loneliness commonly associated with aging.^6^

The walkability of a neighborhood is determined by several factors, including street connectivity and density^7,8^, access to destinations and aesthetics^9,10^, investment in walking and biking infrastructure^11^, and the presence/absence of urban natural features, specifically tree cover.^12^ Several recent studies have investigated the effects of greenspace on physical activity, providing consistent evidence of the beneficial effects of urban nature-based infrastructure on health outcomes.^13,14^ Tree canopy strongly influences urban air temperature^12^ and can reduce land surface temperature by as much as 2.9°F.^15^

Despite the growing evidence of the cooling effect of vegetation and tree cover in urbanized areas, many neighborhoods in impoverished and minority communities have been found to lack access to these ecosystem services.^12^ The patterns of inequality in greenspace distribution and associations with hotter microclimates emerge in multiple incorporated and unincorporated urban areas across the United States.^12,16–21^ These patterns contribute to the urban heat island effect first described by Oke *et al.* in a study of sensible heat storage in the urban core of Mexico City.^22^ In essence, the urban heat island effect is the persistence of higher temperatures in urban centers compared to the surrounding rural or natural areas.^23^ Land surface temperature (LST) derived from Landsat 8 Thermal Infrared Sensors (TIRS) has been extensively used to demarcate the spatial boundaries of the urban heat island effect.^24–26^

Various studies have assessed the associations between urban heat island effects and exposure of vulnerable populations to heat-related health risks.^16,27–29^ For instance, an in-depth analysis of the causes of the high mortality rates of the 1995 Chicago heat wave study found that disproportionate exposure of socially vulnerable groups, social isolation of seniors, and institutional neglect of poor and racialized neighborhoods have contributed to the observed disastrous outcomes.^27^ A study of the urban heat island effect in 25 cities around the world observed that low-income neighborhoods consistently exhibit lower vegetation density and are more likely to experience higher levels of heat exposure.^18^ Mitchell & Chakraborty^16^ developed a composite “urban heat risk index” based on the physical characteristics of the urban environment, such as LST, urban form and building density, and amount of greenspace. The index was applied to the cities of Los Angeles, Chicago, and New York in conjunction with indicators of social vulnerability. The study found that census tracts with higher percentages of racial and ethnic minorities had a strong positive correlation with heat exposure, but did not establish consistent evidence of an association between the urban heat risk index and disproportionate exposure of older adults.^16^ Similarly, another study^23^ observed a nonsignificant statistical association between heat exposure and isolation of older adults (percent 65 and older who live alone) in Portland, Oregon. A study that analyzed urban heat islands in 175 large US cities observed that racial and ethnic minorities (versus white) experience higher temperatures in 97% of the studied cases, but found no evidence of higher UHI intensities for older adults.^21^ Using remote sensing and census data, another study established widespread urban heat disparities in 1000 counties in the continental US.^20^ Lastly, a study that explored the association between tree cover, land surface temperatures, and income in 5,723 US communities found that poorer neighborhoods have, on average, 15% less vegetative cover and are more likely to experience higher temperatures (as much as 1.5°C hotter) than more affluent subdivisions.^12^ Moreover, the study established a strong correlation between a higher percentage of tree cover and higher income in 92% of the urbanized areas.^12^

Research suggests that historic policies of housing discrimination, such as redlining, have contributed to the disproportionate exposure of low-income urban populations to extreme heat.^14,28–30^ Redlining, instituted in the US in the 1930s, was a lending practice that discriminated against individuals living in minority, low-income, and migrant communities.^28–31^ These neighborhoods were redlined or color-coded red (“hazardous”) on maps used for mortgage/insurance considerations by the Home Owners’ Loan Corporation (HOLC), resulting in the denial of mortgage and property insurance applications.^28–30^ One potential impact of historic redlining policies is the lasting impact on the built environment^31^, including less greenspace/tree canopy and more pavement, leading to an increased chance of urban heat island effects. A study examining intra-urban LST anomalies in 108 urbanized areas in the US found that historically redlined neighborhoods, which remain predominantly communities of color, are experiencing higher levels of intra-urban heat than their more affluent counterparts.^28^ Another study examined LST derived from remote sensing (as a measure of heat exposure) and the Normalized Difference Vegetation Index (NDVI) (as a measure of vegetative cover) in formerly redlined neighborhoods in Baltimore, Dallas, and Kansas City.^29^ The findings suggest that mean NDVI was lower while mean LST was higher in nearly all historically redlined neighborhoods compared to higher-income communities. An investigation of heat-related emergency room visits in 11 Texas cities found a significant association between a greater percentage of redlined areas, elevated levels of heat exposure, and higher rates of outpatient visits and inpatient admissions.^30^ Lastly, individuals ≥65 years old in the 2017 National Household Travel Survey who lived in historically redlined communities reported spending less time walking in their neighborhoods.

Despite prior evidence suggesting that various health outcomes are affected by historic redlining and heat exposure, few studies have examined how redlining and urban heat islands may impact neighborhood walking in older adults. To address these gaps, in this study we assessed: 1) how individual and neighborhood characteristics differ (including historic redlining score) depending on neighborhood maximum summer LST; 2) whether higher LST is associated with less neighborhood walking among older adults, and whether these associations differ by historic redlining score; and 3) whether neighborhoods with discriminatory historic redlining scores have greater LSTs.

## Methods

We used data from the 2017 National Household Travel Survey (NHTS), a cross-sectional survey of travel behavior (e.g., travel modes and destinations) completed by 264,234 individuals from 129,696 households. NHTS respondents completed online/phone surveys and recorded trip details during a single travel diary day (Monday-Sunday). The study was deemed not human subjects research by University of Miami’s Institutional Review Board. We restricted it to ≥ 65-year-olds living in US Census tracts with data available on historical redlining scores and summer LSTs.

### Land surface temperature

Aggregated daytime maximum LST data for a 40-day timespan from July to August 2013 were downloaded from the Center for International Earth Science Information Network. Descriptive statistics, including mean, min, max, range and standard deviation (SD) for LST, were derived for both census tracts and core-based statistical areas (CBSA) using the zonal statistics algorithm in ESRI’s ArcMap 10.8.1. Standardized scores were used to compare LST values at the respondent’s Census tract level to the surrounding region/CBSA and represented the standardized peak summertime LST. The LST z-scores used in our analyses were calculated by dividing the difference between the mean LSTs at the census tract and CBSA levels by the SD of the LST for the CBSA. More positive z-scores indicate that the participant’s neighborhood (Census tract) had a higher maximum LST than the surrounding region.

### Neighborhood walking

Self-reported time on walking trips was obtained from travel diaries that were completed by NHTS participants on their assigned diary day. Any walking trip that began, ended, or originated and terminated at the respondent’s residence was included in the calculation of total minutes spent walking in the neighborhood for that day.

### Neighborhood characteristics

We linked data on social and built environment characteristics, including redlining scores using the respondents’ US Census tract IDs provided by Federal Highway Administration. HOLC map mortgage investment risk scores were downloaded from a publicly available website (scores: 1=” best”, 2=” still desirable”, 3=” definitely declining”, and 4=” hazardous”). Meier et al^32^ previously derived these scores using equal intervals to assign the four redlining score categories to each US Census tract.

Population density (people/mi^2^) and percentage of African American/Black and Hispanic/LatinX residents at the census tract level were obtained from the NHTS dataset and the American Community Survey (ACS, 2017 5-year estimates). The area deprivation index (ADI), downloaded from University of Wisconsin’s Neighborhood Atlas, ranks census block groups by neighborhood disadvantage based on 2011-2015 ACS estimates of income, education, employment, and housing (1%-lowest to 100%-highest disadvantage).^33^ We derived a tract-level measure by averaging the block-group ADI values within respondents’ Census tracts. We downloaded 2011 National Land Cover Dataset (NLCD) data from the National Historic GIS website (NHGIS).^34^ NHGIS processed NLCD data to determine the proportion of each tract covered by 16 land cover types (e.g., high intensity development, developed open space, forest). For this study, we calculated the percentage of the respondent’s census tract composed of any greenspace (i.e., developed open space, forest, shrub/scrub; grassland/herbaceous, farmland, and wetlands). Further information on the NLCD classifications is published elsewhere ^35,36^. We also report the participants’ Census regions as walking differences have been observed among older adults based on Census region.^37^

### Covariates

The respondent demographics included age (years), sex, education (<high school degree, high school degree, some college, bachelor’s degree, graduate/professional degree), 4-category household income (originally collected in $10,000/year categories), race/ethnicity (non-Hispanic White, African American/Black, Hispanic, Asian, other).

### Statistical methods

We calculated frequencies and percentages for the respondent demographics and neighborhood built and social environment characteristics (including redlining score) by neighborhood LST z-score quartile (Q1=coolest; Q4=hottest). Mean minutes of neighborhood walking per day was calculated by redlining score and LST z-score quartile. We tested differences in sample characteristics and minutes of neighborhood walking by LST quartile and redlining score using unadjusted ordinal logistic regression and linear regression.

Multivariable negative binomial regression models, accounting for clustering of participants within households and within census tracts, tested associations between LST z-score and neighborhood walking minutes/day. Negative binomial regression accounted for zero inflated/skewed data (model fit confirmed via Pearson X^2^ goodness-of-fit test). The models controlled for age, sex, education, income, race/ethnicity, and state of residence. Regression estimates were exponentiated to determine prevalence ratios and 95% confidence intervals (CI). The models were repeated after stratifying by historic redlining score (“definitely declining” or “hazardous” versus “still desirable” or “best”) to assess whether individuals in neighborhoods with both discriminatory historic redlining scores and greater LST demonstrate the least amount of neighborhood walking/day. Lastly, a linear regression model that controlled for state of residence and used generalized estimating equations to account for clustering by household tested the association between living in a neighborhood with a redline score of “definitely declining” or “hazardous” (versus “still desirable” or “best”) and LST z-score. All analyses were conducted in SAS v9.4.

## Results

The analytic sample (n=3,982) was on average 73 years old (SD=7.2), 55.1% were women, and 70.2% had at least some college education (Table 1). Seventy-eight percent were non-Hispanic White, 10% were African American/Black, 7% were Hispanic, 3% were Asian, and 2% were of another race/ethnicity. Hispanic and Asian individuals and those with annual household incomes >$125,000/year were less frequently living in neighborhoods with higher peak summer LSTs.

**Table 1.**
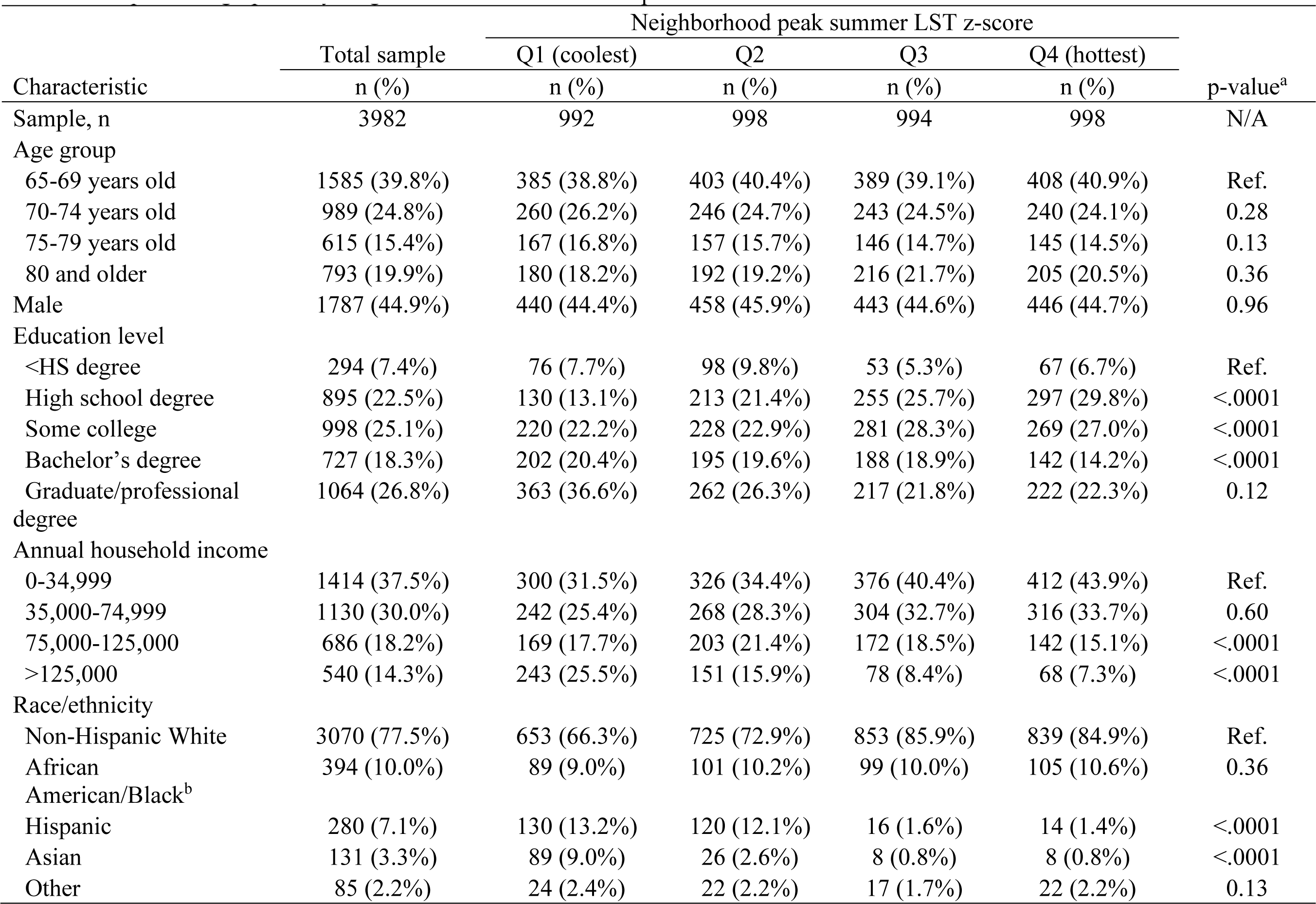
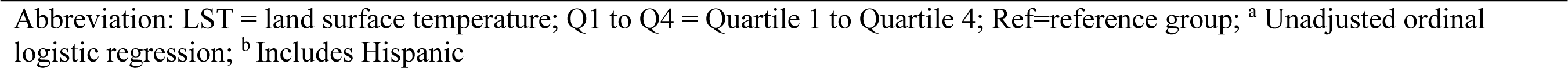
Sample demographics by neighborhood land surface temperature.

Figure 1 depicts the variation in peak summer LST across the US in 2013. Individuals living in the South and West but not the Northeast and Midwest were more likely to live in neighborhoods with lower peak summer LSTs (Table 2). Individuals in neighborhoods with higher area deprivation index scores and greater neighborhood percentage of African American/Black residents were more likely to experience higher peak summer LSTs. Individuals living in neighborhoods with redlining scores of “best” were more likely to live in areas with lower peak summer LSTs.

**Figure 1.**
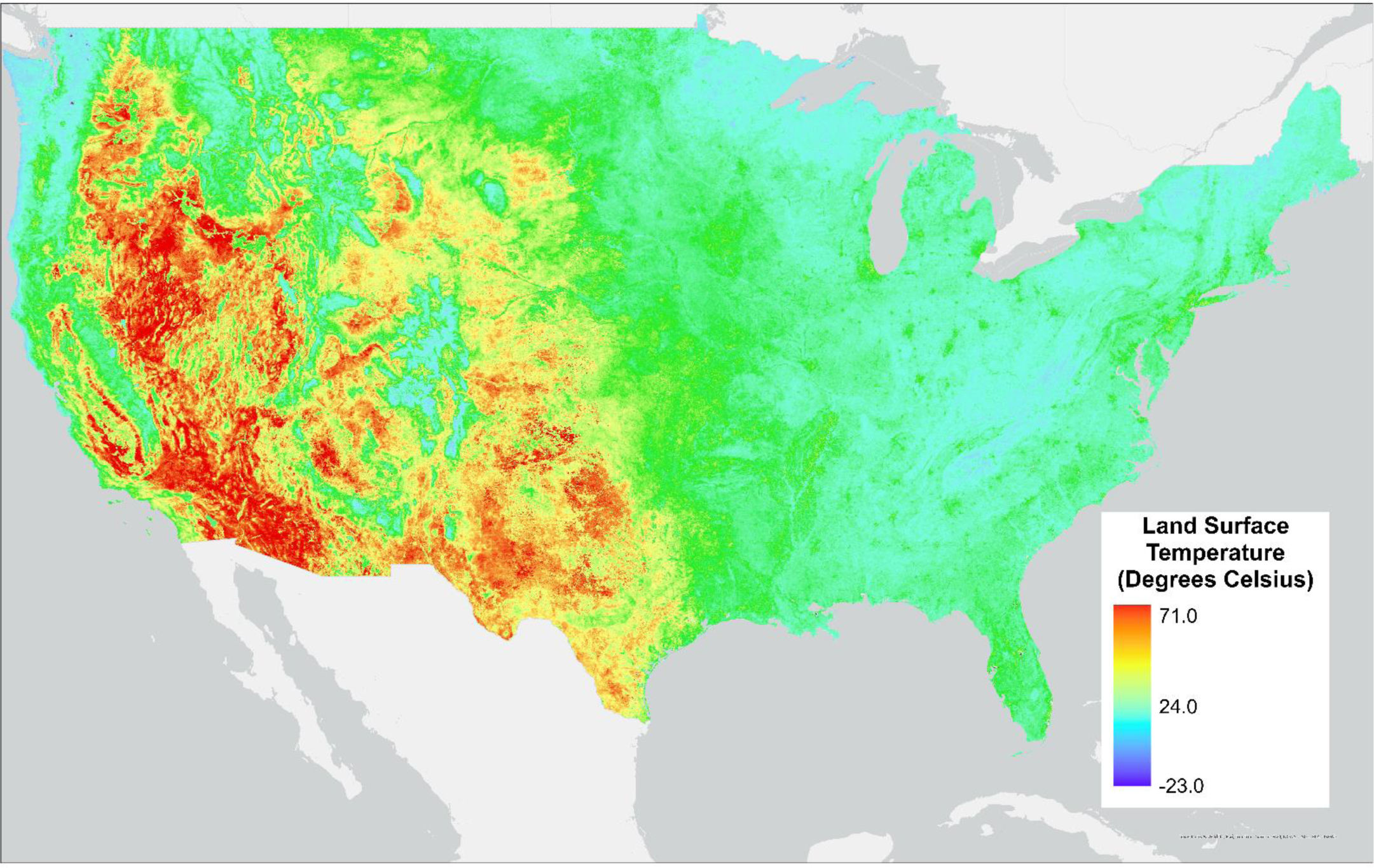
Averages Land Surface temperature for a 40-day timespan from July to August 2013 (*Data Source*: Center for International Earth Science Information Network (CIESIN)

**Table 2.**
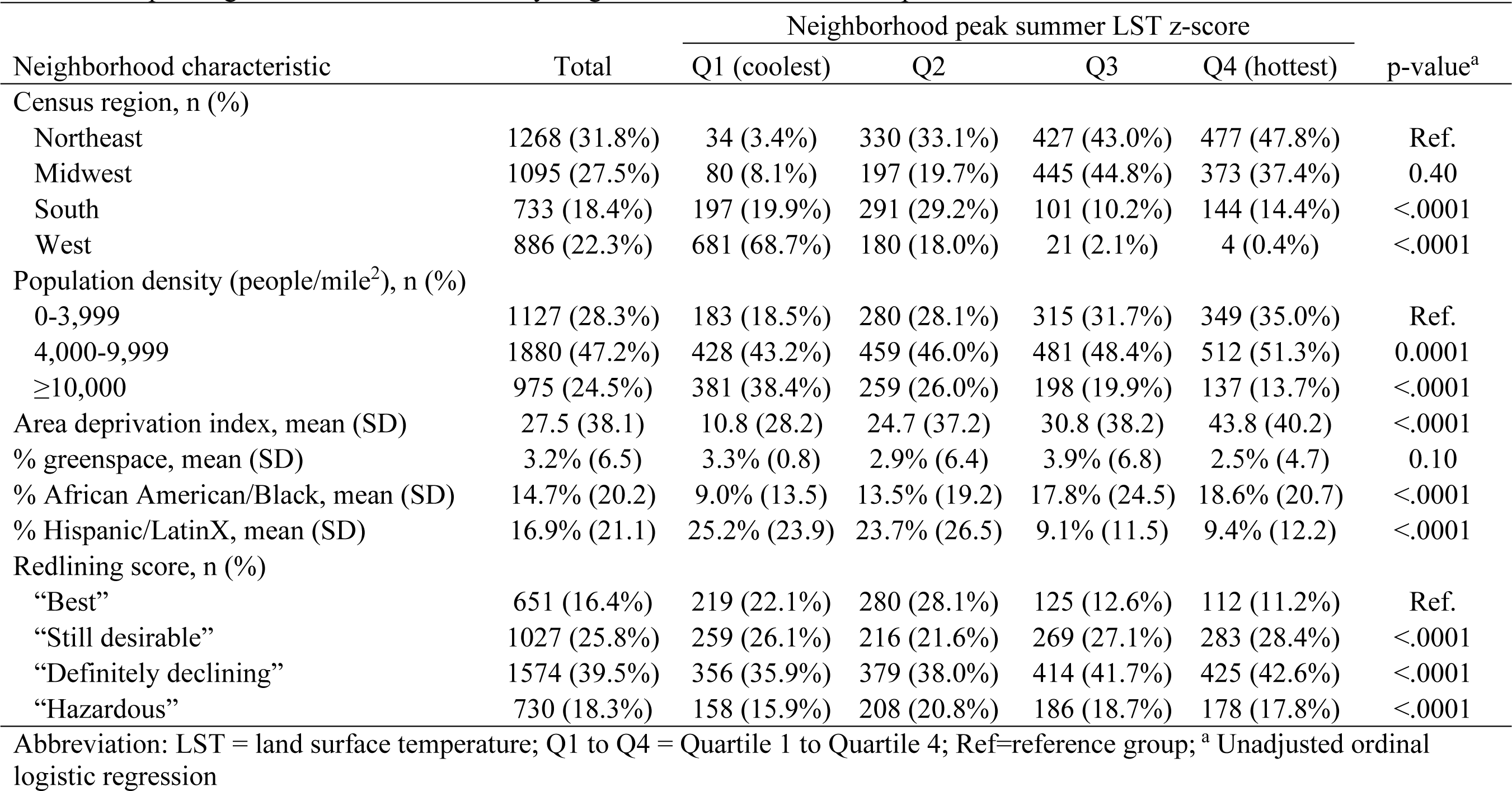
Sample neighborhood characteristics by neighborhood land surface temperature.

| Neighborhood characteristic | Total | Neighborhood peak summer LST z-score |  |  |  | p-value<sup>a</sup> |
| --- | --- | --- | --- | --- | --- | --- |
|  |  | Q1 (coolest) | Q2 | Q3 | Q4 (hottest) |  |
| Census region, n (%) |  |  |  |  |  |  |
| Northeast | 1268 (31.8%) | 34 (3.4%) | 330 (33.1%) | 427 (43.0%) | 477 (47.8%) | Ref. |
| Midwest | 1095 (27.5%) | 80 (8.1%) | 197 (19.7%) | 445 (44.8%) | 373 (37.4%) | 0.40 |
| South | 733 (18.4%) | 197 (19.9%) | 291 (29.2%) | 101 (10.2%) | 144 (14.4%) | <.0001 |
| West | 886 (22.3%) | 681 (68.7%) | 180 (18.0%) | 21 (2.1%) | 4 (0.4%) | <.0001 |
| Population density (people/mile<sup>2</sup>), n (%) |  |  |  |  |  |  |
| 0-3,999 | 1127 (28.3%) | 183 (18.5%) | 280 (28.1%) | 315 (31.7%) | 349 (35.0%) | Ref. |
| 4,000-9,999 | 1880 (47.2%) | 428 (43.2%) | 459 (46.0%) | 481 (48.4%) | 512 (51.3%) | 0.0001 |
| ≥10,000 | 975 (24.5%) | 381 (38.4%) | 259 (26.0%) | 198 (19.9%) | 137 (13.7%) | <.0001 |
| Area deprivation index, mean (SD) | 27.5 (38.1) | 10.8 (28.2) | 24.7 (37.2) | 30.8 (38.2) | 43.8 (40.2) | <.0001 |
| % greenspace, mean (SD) | 3.2% (6.5) | 3.3% (0.8) | 2.9% (6.4) | 3.9% (6.8) | 2.5% (4.7) | 0.10 |
| % African American/Black, mean (SD) | 14.7% (20.2) | 9.0% (13.5) | 13.5% (19.2) | 17.8% (24.5) | 18.6% (20.7) | <.0001 |
| % Hispanic/LatinX, mean (SD) | 16.9% (21.1) | 25.2% (23.9) | 23.7% (26.5) | 9.1% (11.5) | 9.4% (12.2) | <.0001 |
| Redlining score, n (%) |  |  |  |  |  |  |
| “Best” | 651 (16.4%) | 219 (22.1%) | 280 (28.1%) | 125 (12.6%) | 112 (11.2%) | Ref. |
| “Still desirable” | 1027 (25.8%) | 259 (26.1%) | 216 (21.6%) | 269 (27.1%) | 283 (28.4%) | <.0001 |
| “Definitely declining” | 1574 (39.5%) | 356 (35.9%) | 379 (38.0%) | 414 (41.7%) | 425 (42.6%) | <.0001 |
| “Hazardous” | 730 (18.3%) | 158 (15.9%) | 208 (20.8%) | 186 (18.7%) | 178 (17.8%) | <.0001 |

Abbreviation: LST = land surface temperature; Q1 to Q4 = Quartile 1 to Quartile 4; Ref=reference group; <sup>a</sup> Unadjusted ordinal logistic regression

Older adults living in neighborhoods with historic redlining scores of “still desirable”, “definitely declining” or “hazardous” reported fewer minutes of neighborhood walking/day compared to those in neighborhoods with historic redlining scores of “best” (Table 3). Compared to individuals living in neighborhoods at the bottom quartile of peak summer LSTs, those in the top quartile walked significantly fewer minutes in the neighborhood per day.

**Table 3.**
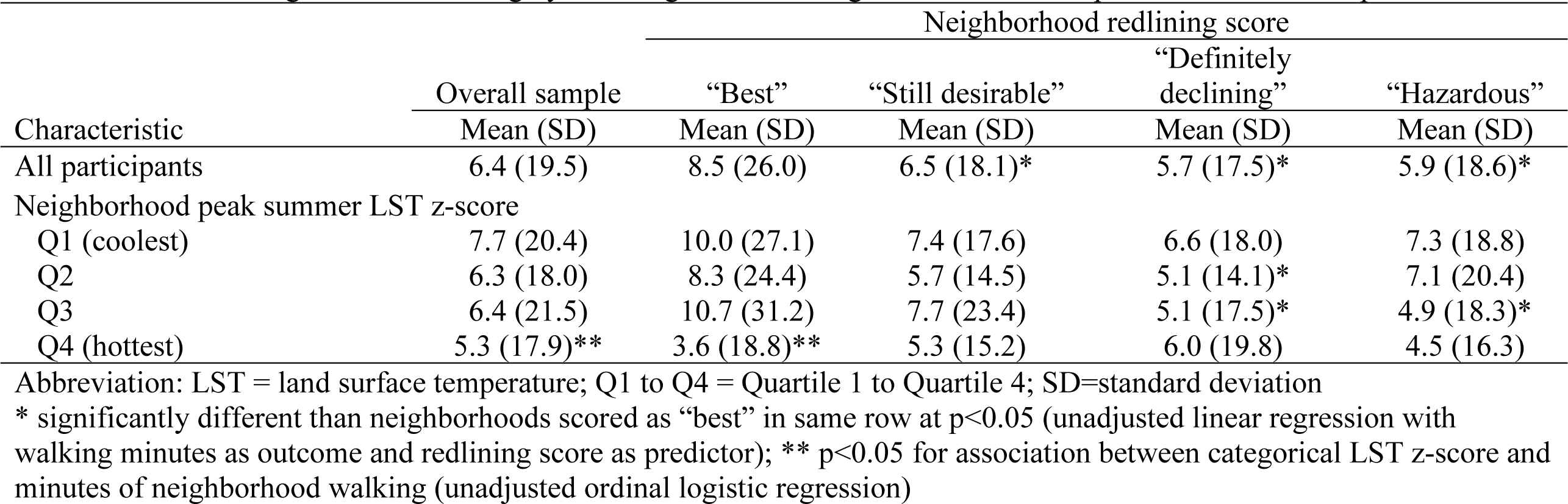
Minutes of neighborhood walking by redlining score and neighborhood summer peak land surface temperature.

In multivariable regression analyses, older adults living in neighborhoods with higher peak summer LST z-scores reported fewer minutes of neighborhood walking/day (Table 4) (PR: 0.85, 95% CI: 0.74-0.98). Stratifying by redlining score showed no significant associations between LST z-score and neighborhood walking in separately for those living in neighborhoods with redlining scores of “definitely declining” or “hazardous” (PR: 0.99, 95% CI: 0.82-1.21) or for those living in neighborhoods scored as “still desirable” and “best” (PR: 0.77, 95% CI: 0.58-1.01). However, the latter association was borderline significant at p=0.059. Lastly, neighborhood redlining scores of “definitely declining” or “hazardous” (versus “still desirable” and “best”) were associated with greater neighborhood peak LSTs (Table 5) (estimate: 0.103, 95% CI: 0.009-0.198).

**Table 4.** Association between neighborhood summer peak land surface temperature and minutes of neighborhood walking per day.

| Model | PR<sup>a</sup> | (95% CI) | p-value |
| --- | --- | --- | --- |
| Overall sample | 0.85 | 0.74-0.98 | 0.02 |
| Neighborhood had historic redlining score of “Definitely Declining” or “Hazardous” | 0.99 | 0.82-1.21 | 0.95 |
| Neighborhood had historic redlining score of “Still Desirable” or “Best” | 0.77 | 0.58-1.01 | 0.059 |

<sup>a</sup> Controlling for age, sex, education, income, race/ethnicity, state

**Table 5.** Association between dichotomous neighborhood redlining score and Neighborhood peak summer land surface temperature.

| Variable | Estimate (95% CI)<sup>a</sup> | p-value |
| --- | --- | --- |
| Neighborhood scored as “definitely declining” or hazardous”<sup>b</sup> | 0.103 (0.009-0.198) | 0.03 |

<sup>a</sup> Controlled for state of residence
<sup>b</sup> versus “still desirable” or “best”

## Discussion

The number of empirical studies on the relationship between the historical policies of housing discrimination, urban greenspace availability, and exposure to urban heat anomalies in economically disadvantaged communities of color continues to grow, revealing consistent patterns of inequality.^28–31^ However, despite the attention given to the urban heat island effect and its implications for human health and well-being in recent years, relatively few studies have specifically addressed the exposure of older adults to peak summer LSTs in historically redlined neighborhoods and the extent to which this exposure affects daily physical activity. This study attempts to fill these knowledge gaps by leveraging data from travel diaries on daily walking trips, peak summer LSTs, neighborhood characteristics, and the historical legacy of residential segregation to assess the physical activity patterns of 3,982 adults 65 years and older.

We found evidence of significant peak summer warming in census tracts with higher area deprivation index scores. Heat exposure variation was also observed in relation to the neighborhoods’ racial and ethnic composition. A higher heat risk burden was associated with communities characterized by a larger percentage of African American/Black residents. We found that lower-income, Asian, and Hispanic individuals were more likely to live in neighborhoods with higher peak summertime LST compared to White and higher-income residents. These outcomes corroborate the results of previous studies, which consistently found evidence of urban heat exposure disparities associated with income and race/ethnicity.^20,28^

Several recent studies highlighted the legacy of institutionalized residential segregation after the Great Depression and the impact of systematic targeting of communities of color, which ultimately led to disinvestment and profound socio-economic inequalities.^29,30^ We observed a greater neighborhood peak summertime LSTs in neighborhoods with redlining scores of “definitely declining” or “hazardous” (versus “still desirable” and “best”). Our findings confirm the results from previous studies. For example, Li et al. observed that the proportion of redlined areas is a significant predictor of both daytime and nighttime LST.^30^ We also found that older adults reported fewer minutes of walking in historically redlined neighborhoods with Home Owners’ Loan Corporation (HOLC) scores of “definitely declining” and “hazardous.”

Several limitations of this study warrant further consideration. Landsat data has a repeat interval of approximately two weeks (with no more than two observations in any given month). Extensive cloud cover, typical for wet summer months in certain regions, may obstruct the usability of Landsat data for extended periods of time.^29^ To address this limitation, this study used aggregated daytime maximum LST data for a 40-day timespan from July to August 2013. Previous studies have also highlighted the fact that LST, as a proxy of thermal exposure, may not fully capture the heat stress experienced by individuals.^18,30^ These limitations suggest that more complex modeling of urban microclimates that takes into account both LST and air temperature, as well as the cooling effects of urban blue-green infrastructure, may improve our understanding of urban heat exposure.

Our results suggest that the association between higher neighborhood LSTs and less neighborhood walking might be restricted to older adults living in neighborhoods with historic redlining scores of “still desirable” or “best” (i.e., advantaged communities). Prior work has demonstrated that historically redlined neighborhoods remain more economically deprived, less walkable, have more less greenspace and tree canopy, and have greater concentrations of African American/Black and Hispanic/LatinX residents.^31^ Conversely, neighborhoods with historic redlining scores of “still desirable” and “best” have built and social environments today that are more conducive to neighborhood walking (i.e., higher SES, more greenspace, more pedestrian amenities and destinations). Yet, we found that individuals in those more affluent neighborhoods walk less if their neighborhoods experience higher LSTs. This may be explained by the lack of transportation options and other sources of recreational physical activity for older adults living in more disadvantaged communities that were historically redlined, and thus their neighborhood walking does not decrease with higher neighborhood LSTs. Older adults in more affluent communities are more likely to have alternative sources of transportation to nearby destinations other than walking, as well as other places and opportunities for exercise other than neighborhood walking ^38^, and thus may be more likely to decrease neighborhood walking with increasing temperatures. If these findings are replicated in other studies, this suggests an even more pressing need to address urban heat islands in historically disadvantaged communities because older adults (who are more susceptible to heat) in these communities are more likely to be exposed to extreme heat in their neighborhoods and experience negative health effects (e.g., heat stroke and cardiorespiratory symptoms).^39^

## Conclusion

Overall, this study suggests that historically redlined neighborhoods may more often experience urban heat island effects and that older adults living in hotter neighborhoods may less often engage in neighborhood walking. Discriminatory policies such as historic redlining have left indelible marks on neighborhoods across the US, leaving communities more vulnerable to detrimental environmental exposures, including extreme heat. Future work is needed to elucidate the impact of extreme heat on health-promoting behaviors such as neighborhood walking and the types of interventions and policies that can successfully counteract the negative health impacts of extreme heat on these historically disadvantaged communities.

## Data Availability

Data are available from the original data source (US Department of Transportation) and publicly available data for download on the National Household Travel Survey website.

https://nhts.ornl.gov/

